# A STUDY ON ASSESSMENT OF MEDICATION ADHERENCE IN HEMODIALYSIS PATIENTS

**DOI:** 10.1101/2025.02.10.25321945

**Authors:** Razieh Bahrami Gahrouei, Srinivasan Ranganathan

## Abstract

**Background:** Medication adherence is a crucial factor in managing chronic diseases such as chronic kidney disease (CKD), particularly in hemodialysis patients who require complex and lifelong medication regimens. Non-adherence in this population is associated with increased morbidity, hospitalizations, and reduced treatment efficacy. This study aims to assess medication adherence levels, identify key demographic and clinical factors influencing compliance, and evaluate the impact of counseling interventions on adherence behavior.

**Methods:** A six-month prospective observational study was conducted at BGS Global Hospital, Bangalore, involving 89 hemodialysis patients (61 males, 28 females). The Morisky Green Levine Scale (MGLS) was used to assess medication adherence before and after structured counseling sessions. Participants were stratified by gender and age groups (18–35, 35–60, and 60–80 years) to analyze adherence patterns. Health-related quality of life (KDQOL) was also evaluated to determine the impact of adherence on patient well-being. Statistical significance was assessed using p-values, with a threshold of p < 0.05 considered significant.

**Results:** Pre-counseling adherence assessments revealed that only 3.37% of participants were highly adherent, while 80.89% had medium adherence and 15.73% exhibited low adherence. Gender analysis showed that males had slightly higher adherence rates before counseling (4.91%) compared to females (0%), while females had a greater proportion of medium adherence (85.71%). Post-counseling, adherence improved significantly, with high adherence increasing to 39.3% in males and 39.2% in females. Age-based analysis indicated that the 35–60 age group demonstrated the highest adherence rates both before and after counseling, while younger patients (18–35 years) had lower baseline adherence but showed significant improvement post-intervention. KDQOL assessments revealed that improved adherence was associated with better quality-of-life scores, particularly among males (p = 0.217 before counseling, 0.256 after) and females (p = 0.060 before, 0.071 after).

**Conclusion:** The findings demonstrate that structured counseling interventions significantly improve medication adherence among hemodialysis patients across gender and age groups. Given the chronic nature of hemodialysis treatment, integrating adherence-enhancing strategies into routine patient care is essential to optimize treatment outcomes and improve quality of life. Future research should focus on tailored adherence interventions, incorporating behavioral strategies, digital health tools, and long-term monitoring approaches to sustain adherence improvements over time.

## Introduction

Medication adherence is a critical determinant of treatment outcomes in chronic diseases (1,2), particularly in patients undergoing long-term therapies such as hemodialysis (3). The World Health Organization (WHO) defines medication adherence as "the degree to which a person’s behavior corresponds with the agreed recommendations from a healthcare provider (4). This encompasses adherence to prescribed medication regimens in terms of dosage, frequency, and duration, which is particularly essential in managing the complex comorbidities associated with chronic kidney disease (CKD) (4,5).

Patients undergoing hemodialysis require complex and multidrug treatment regimens (6), increasing the risk of non-adherence due to factors such as pill burden (7), treatment fatigue (8), and comorbidities (9). Studies have reported medication non-adherence rates ranging from 22% to 74% in hemodialysis patients (10,11), with an average rate of approximately 51%, demonstrating the widespread challenge of compliance in this population (12). Poor adherence to prescribed medications can lead **to** rapid deterioration of kidney function, increased hospitalizations, higher healthcare costs, and an elevated risk of morbidity and mortality(13,14). Thus, addressing medication adherence is a fundamental aspect of optimizing clinical outcomes in hemodialysis patients (15,16).

Adherence behavior is influenced by multiple demographic (17,18), clinical, and psychosocial factors (19), including gender, age, treatment complexity, cognitive function, socioeconomic status, depression, and patient awareness(20,21).

## METHODOLOGY

### Study Design and Setting

This study was designed as a prospective observational study conducted at the Department of Nephrology, BGS Global Hospital, Kengeri, Bengaluru, India. The study was carried out over a period of six months.

### Participant Selection

#### Inclusion Criteria

Patients meeting the following criteria were included in the study:

- Individuals undergoing hemodialysis at the study site.
- Patients of all age groups.
- Both male and female participants.
- Patients with comorbid conditions.

#### Exclusion Criteria

The following patients were excluded from the study:

- Individuals who had undergone multi-organ transplantation.
- Patients with a history of renal transplantation.
- Patients who refused to provide informed consent.

#### Ethical Approval

The study was reviewed and approved by:

1. The Institutional Ethical Committee of PES College of Pharmacy, Bengaluru.
2. The Institutional Ethics Committee of BGS Global Hospital, Bengaluru.

Prior to enrollment, written informed consent was obtained from all participants in accordance with ethical guidelines.

#### Data Collection Instruments

The study utilized the following standardized tools for data collection:

- Informed Consent Form – to document voluntary participation.
- Patient Profile Form / Data Collection Form – to record demographic and clinical data.
- Morisky Green Levine Questionnaire – a validated tool for assessing medication adherence.
- Kidney Disease Quality of Life (KDQOL) Form – to evaluate patients’ quality of life in relation to adherence.

#### Study Procedure

The study was conducted in three phases:

### Stage I – Informed Consent and Patient Education

At the initial visit, the study procedure was explained to all eligible participants in Kannada, Hindi, or English. Patients were given the opportunity to clarify any doubts before providing their signed informed consent. Following consent, patients received education about their medical condition and the importance of medication adherence.

### Stage II – First Follow-up and Data Collection

During the first follow-up, patients were interviewed regarding their medication-taking behavior. The Morisky Green Levine Medication Adherence Scale was administered to assess their adherence level. Data were collected under the supervision of a clinical staff member using the patient profile form. After data collection, basic information about their prescribed medications was provided.

### Stage III – Second Follow-up and Patient Counseling

During the second follow-up, patients received comprehensive counseling on the importance of medication adherence. The session included discussions on the potential complications of non-adherence, strategies to improve compliance, and the benefits of long-term medication adherence.

### Statistical Analysis

All collected data were entered into Microsoft Excel (2016 version) (22–26) and analyzed using SPSS software (version 22) (27–31). Descriptive statistics were used to summarize qualitative data, which were presented as proportions, pie charts, and bar graphs. The Chi-square test was employed to determine statistical significance, with a p-value of <0.05 considered statistically significant.

## RESULTS

### 1. Gender Distribution Among Hemodialysis Patients

The gender distribution of the study population is presented in Table 1 and Figure 1. A total of 89 patients were included in the study, with a higher proportion of males (n = 61, 68.53%) compared to females (n = 28, 31.46%). This disparity may reflect the demographic composition of patients undergoing hemodialysis at the study site.

**Table 1.** Gender Distribution of Study Participants.

| GENDER | NUMBER OF PARTICIPANTS | PERCENTAGE(%) |
| --- | --- | --- |
| MALE | 61 | 68.53 |
| FEMALE | 28 | 31.46 |
| TOTAL | 89 | 100 |

**Figure 1.**
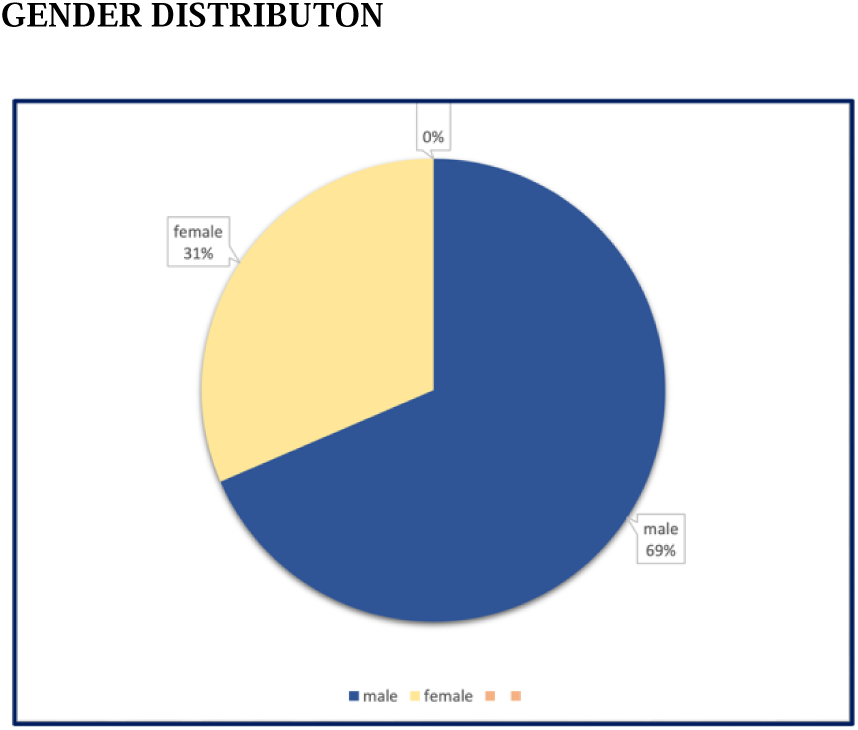
Gender distribution among hemodialysis patients. A total of 89 participants were included in the study, with 61 (68.53%) being male and 28 (31.46%) being female. The figure illustrates the predominance of male patients undergoing hemodialysis at the study site.

### 2. Medication Adherence Levels Among Hemodialysis Patients

Medication adherence among the study participants was assessed using the Morisky Green Levine Scale, which categorizes adherence into high, medium, and low levels based on patient responses. As shown in Table 2, the majority of patients (n = 72, 80.89%) exhibited a medium level of adherence, while 14 patients (15.73%) demonstrated low adherence. Only 3 patients (3.37%) were classified as highly adherent to their prescribed medication regimens.Given that adherence to medication is critical in hemodialysis patients for managing comorbid conditions and improving treatment outcomes, the observed low rate of high adherence suggests a need for enhanced patient education and adherence strategies to improve compliance rates.

### 3. Graphical Representation of Medication Adherence Levels

To further illustrate the distribution of medication adherence among the study population, Figure 3 presents a bar graph depicting the percentage of participants categorized under high, medium, and low adherence levels.The majority of patients (n = 72, 80.89%) exhibited medium adherence, while 14 patients (15.73%) had low adherence, and only 3 patients (3.37%) were classified as highly adherent. These findings highlight the significant proportion of patients who do not fully comply with their prescribed medication regimens, which may impact disease progression and overall treatment efficacy in hemodialysis patients.

**Figure 2.**
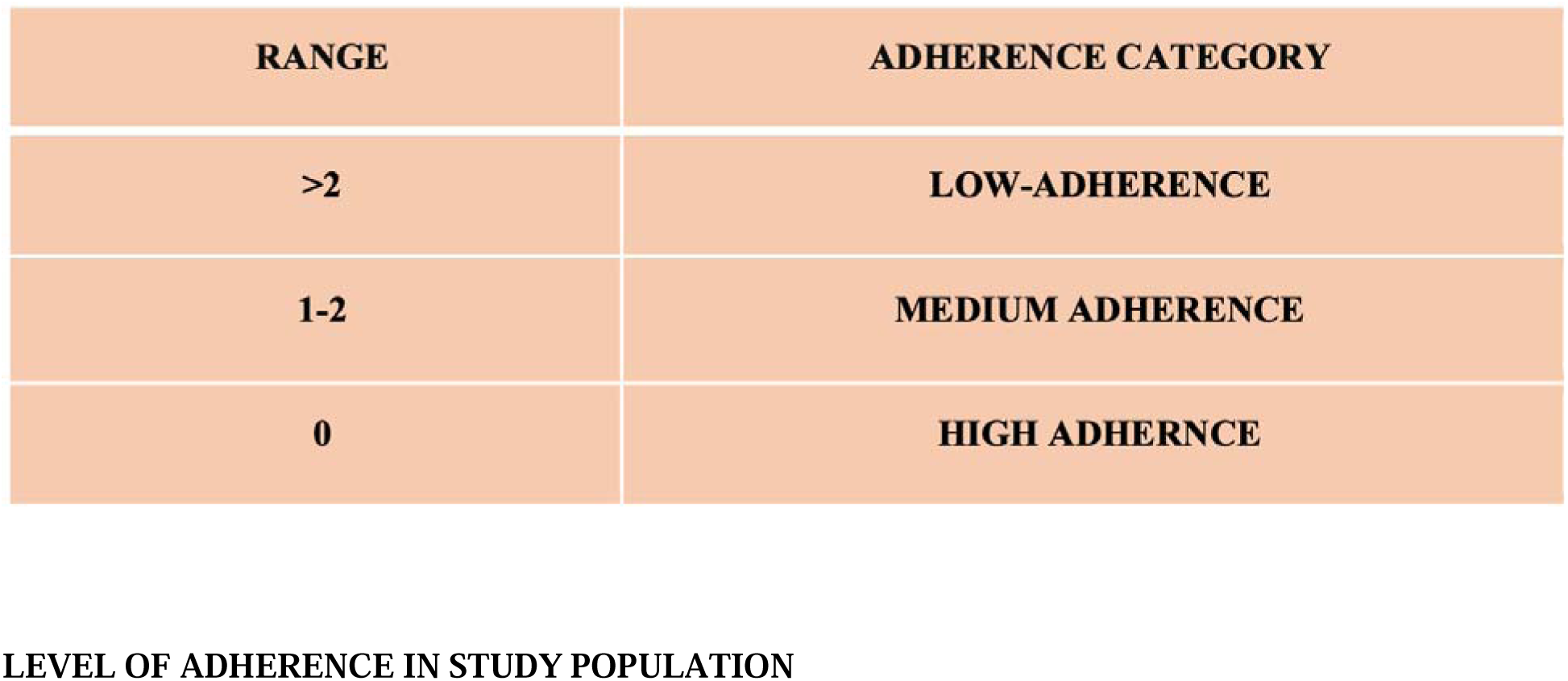
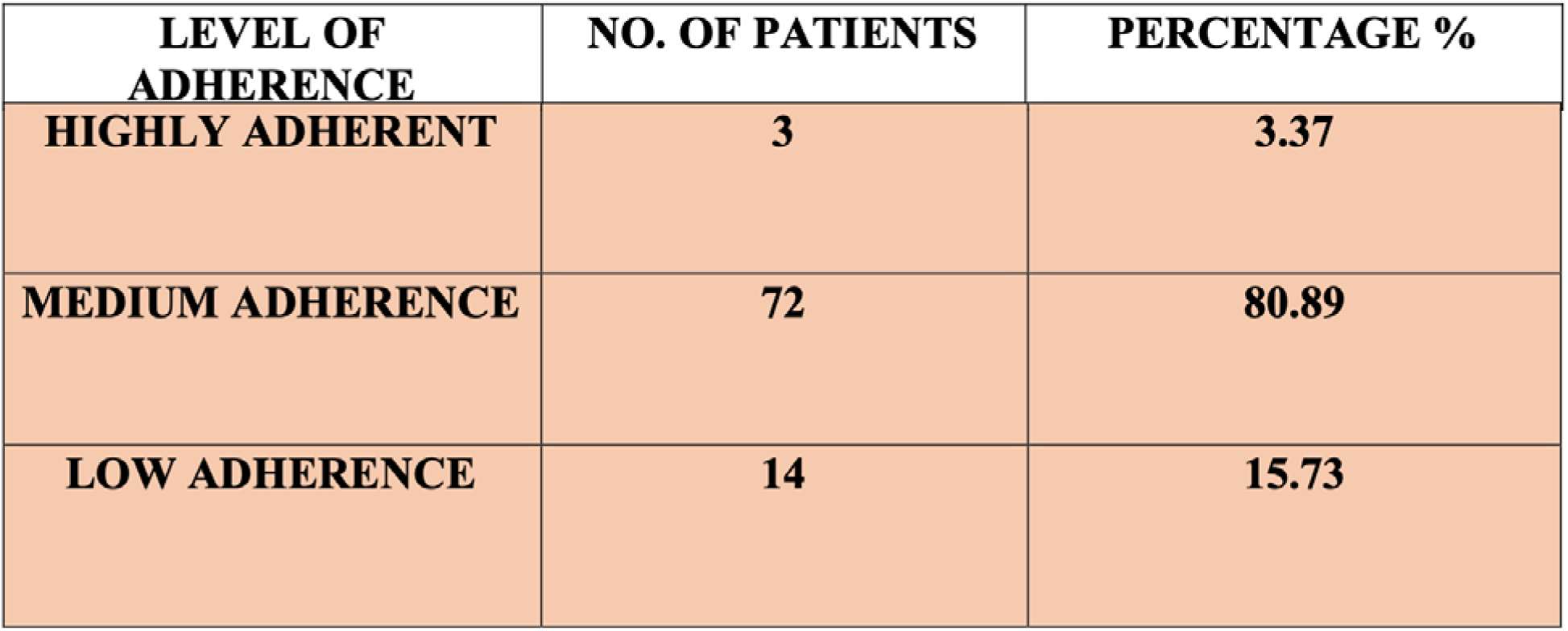
Distribution of medication adherence levels among hemodialysis patients. The figure illustrates the proportion of patients categorized as highly adherent (3.37%), medium adherent (80.89%), and low adherent (15.73%) based on the Morisky Green Levine Scale. The majority of patients exhibited medium adherence, highlighting the need for targeted interventions to improve compliance.

**Figure 3.**
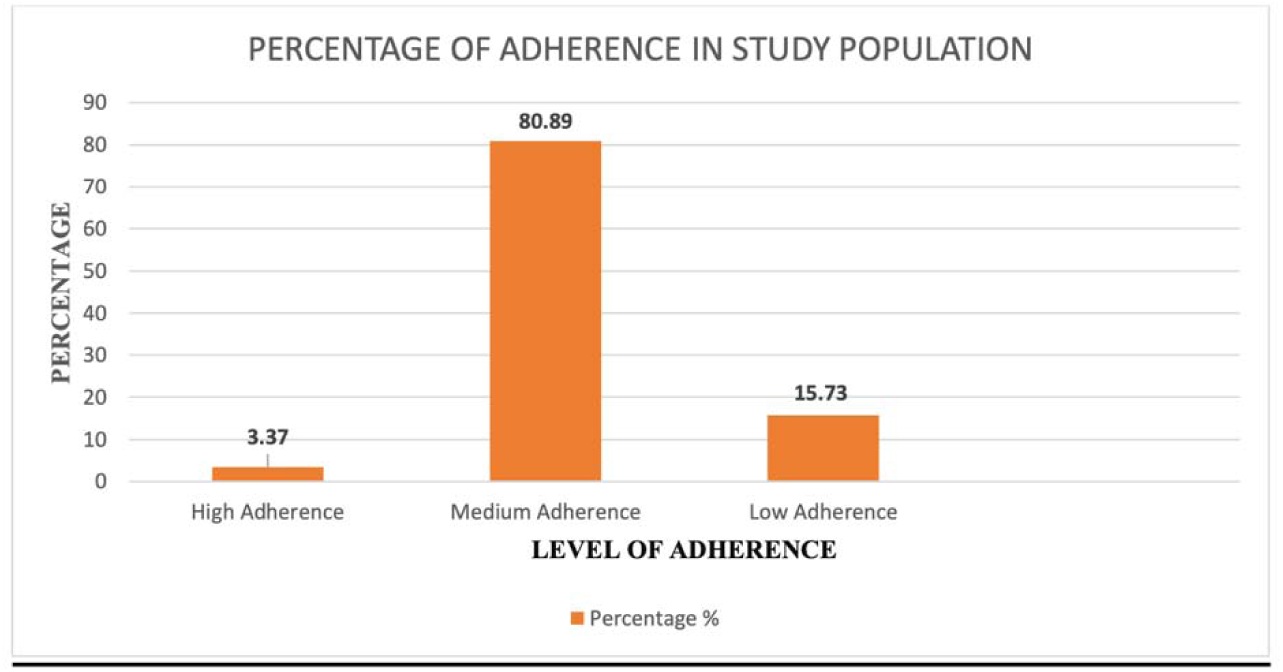
Graphical representation of medication adherence levels in the study population. The bar graph illustrates the percentage distribution of medication adherence levels among the 89 study participants. Medium adherence was the most prevalent (80.89%), followed by low adherence (15.73%), while high adherence was observed in only a small fraction (3.37%) of patients. The findings emphasize the need for enhanced adherence interventions in hemodialysis care.

### 4. Gender-Based Comparison of Medication Adherence Levels

A gender-based analysis of medication adherence was conducted to assess differences in compliance between male and female participants. Table 3 presents the distribution of adherence levels across genders, while Figures 4 and 5 provide graphical representations for male and female participants, respectively.Among male participants (n = 61), 3 (4.91%) exhibited high adherence, while 48 (78.68%) had medium adherence, and 10 (16.39%) demonstrated low adherence. In contrast, among female participants (n = 28), none were highly adherent, while 24 (85.71%) had medium adherence, and 4 (14.28%) showed low adherence.These findings suggest that males had a slightly higher proportion of low adherence compared to females (16.39% vs. 14.28%), while females demonstrated higher medium adherence rates (85.71% vs. 78.68%). The lack of high adherence among female participants highlights a potential area for targeted intervention to improve compliance.

**Figure 4.**
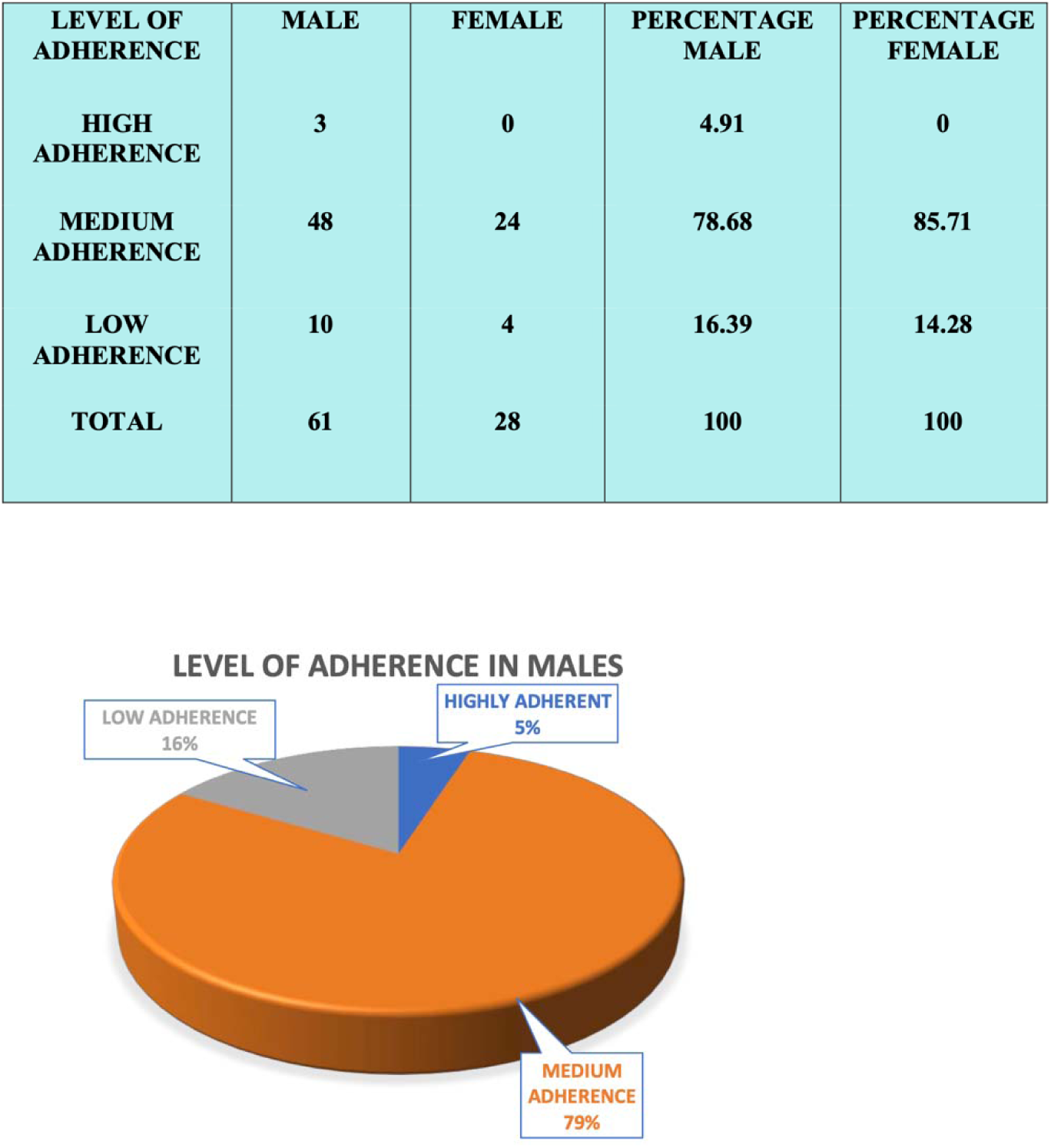
Distribution of medication adherence levels among male participants. The pie chart represents adherence levels among male hemodialysis patients (n = 61). A majority (78.68%) exhibited medium adherence, while 16.39% showed low adherence and only 4.91% were categorized as highly adherent. The pie chart illustrates adherence levels among female hemodialysis patients (n = 28). While most females (85.71%) demonstrated medium adherence, 14.28% showed low adherence, and no participants were categorized as highly adherent.

**Fig5.**
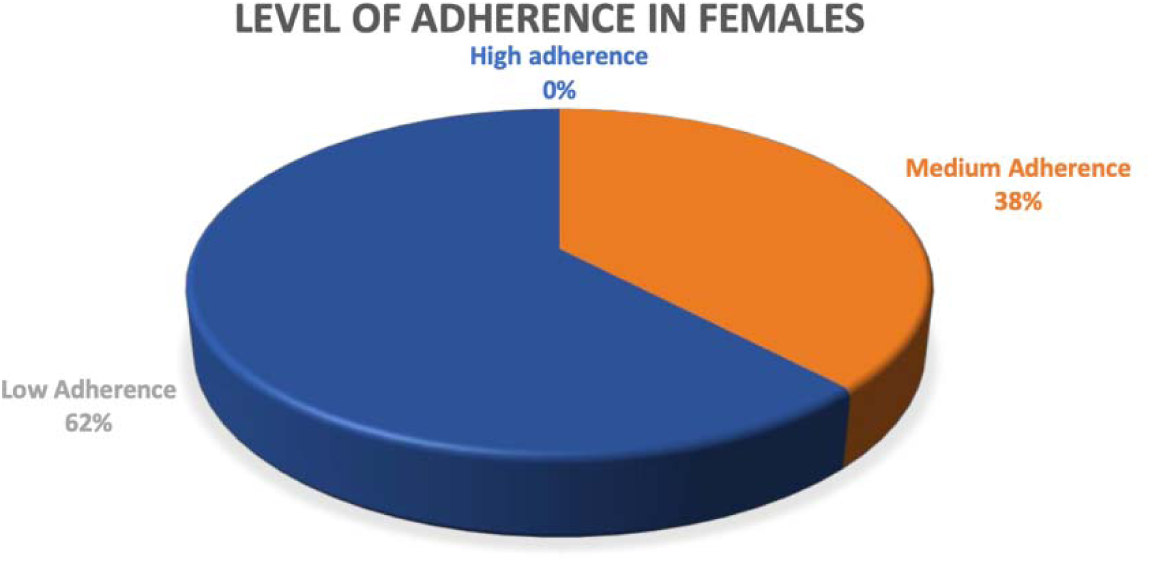
Graphical Representation of level of adherence in females before counselling.

### 5. Gender-Based Medication Adherence Before Counseling

A gender-stratified analysis of medication adherence before counseling was conducted to evaluate differences in adherence behavior between male and female participants. The distribution of adherence levels for male participants (n = 61) is presented in Figure 6, while adherence levels for female participants (n = 28) are depicted in Figure 7.

**Figure 6.**
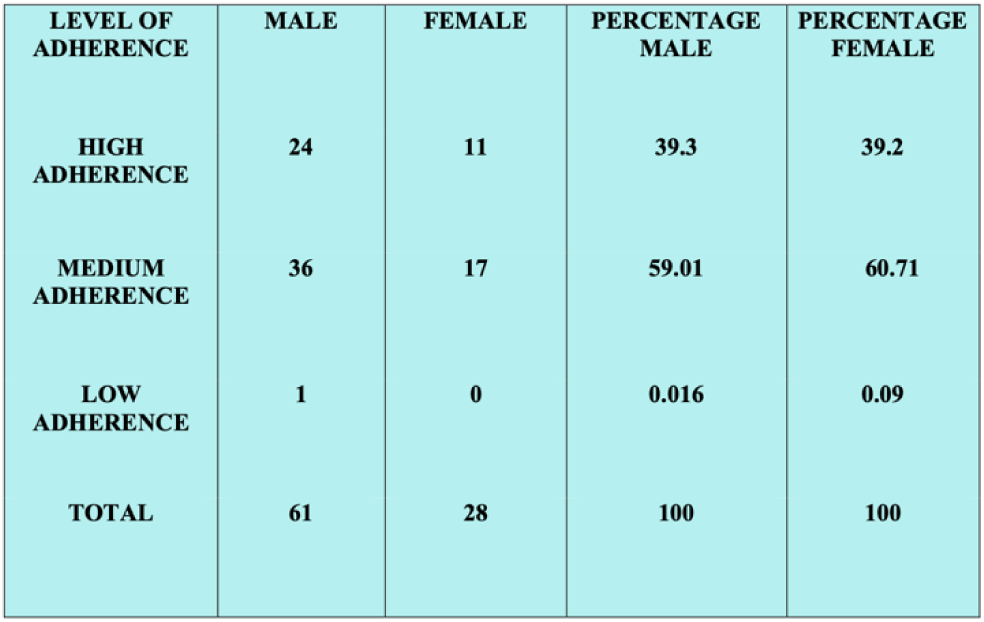
Gender-based distribution of medication adherence after counseling. The table presents the medication adherence levels among male and female hemodialysis patients following counseling interventions. A significant increase in high adherence was observed in both groups, with 39.3% of males and 39.2% of females categorized as highly adherent. The results suggest that targeted counseling improved adherence behavior across genders.

**Fig7.**
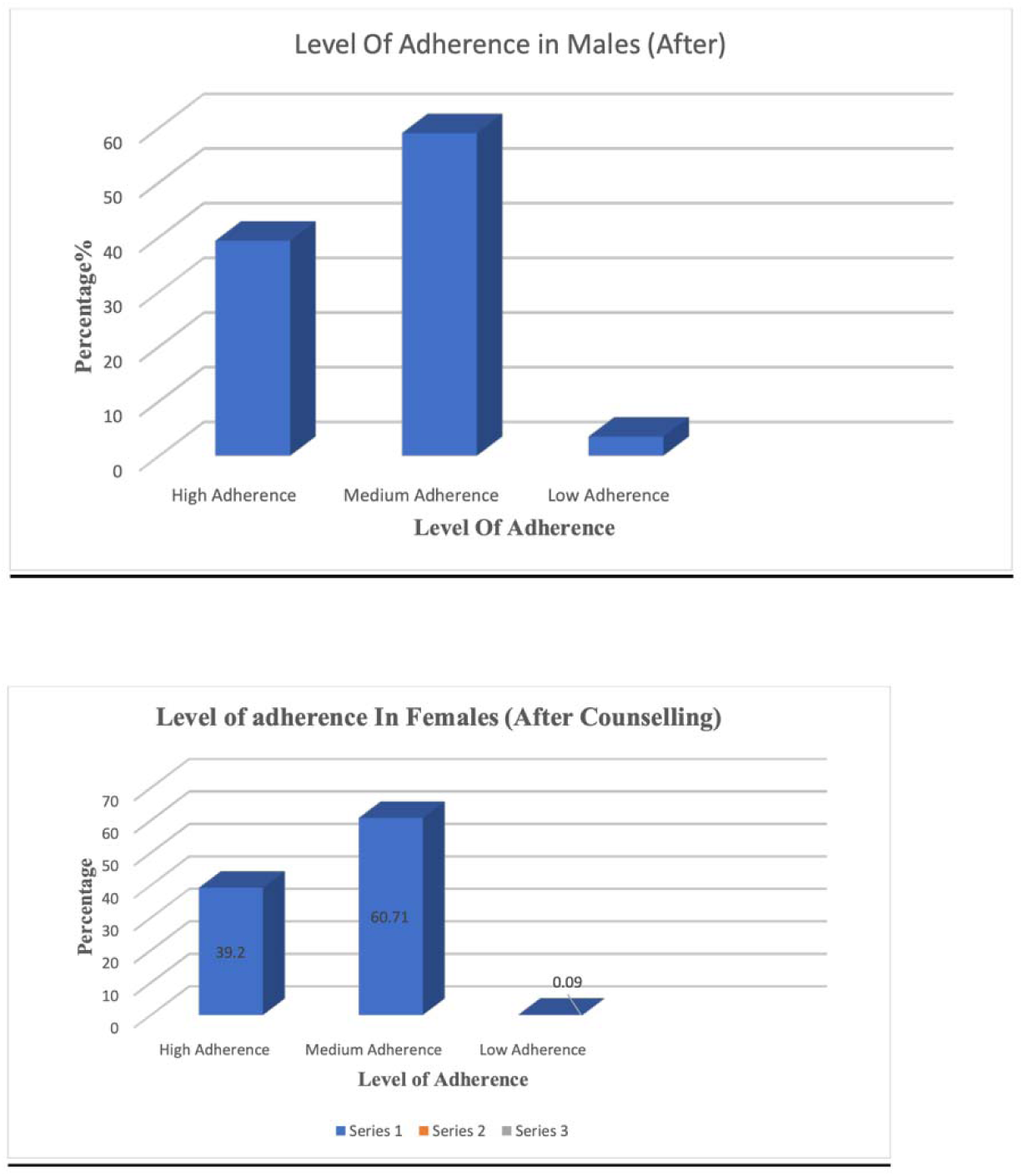
Graphical Representation of Level of Adherence in Females (After counselling) Categorization of adherence based on gender (after counselling) describes that 39.3 % high adherence was found in males and 39.2% was observed in females. Medium level of adherence is seen in 59.01% males and 60.71% females. Low level of adherence was observed in 0.016% and 0% in females. So higher adherence was observed in males than females.

**Figure 8.**
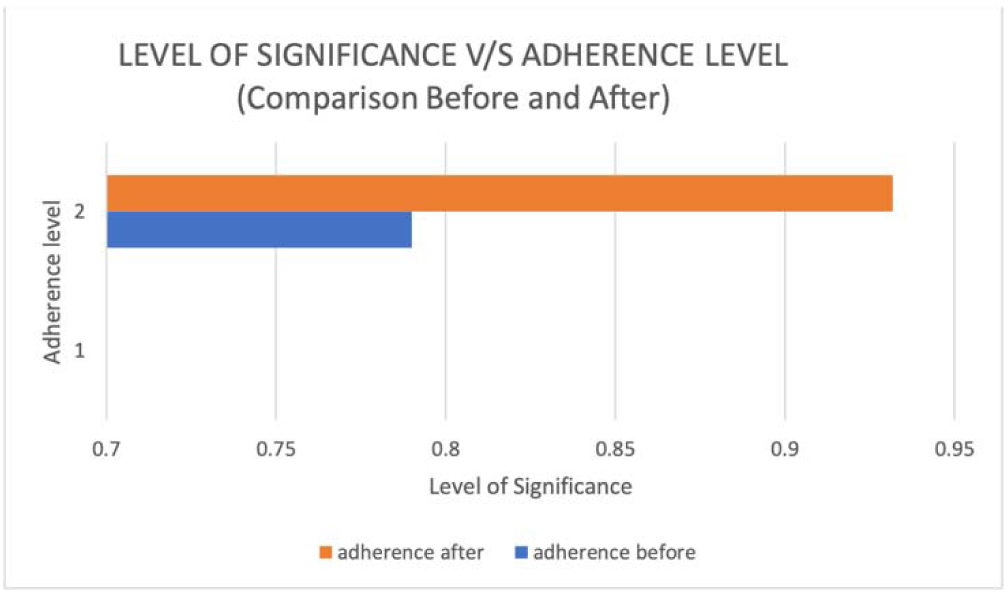
Statistical comparison of adherence levels before and after counseling. The bar chart illustrates the level of significance in medication adherence before and after counseling interventions. The adherence level significantly improved post-counseling, as indicated by the higher significance level in the orange bar. A Chi-square test confirmed statistical significance (p < 0.05), demonstrating the positive impact of counseling on adherence behavior.

Among male participants, only 3 individuals (4.91%) demonstrated high adherence, while 48 (78.68%) exhibited medium adherence, and 10 (16.39%) had low adherence. For female participants, none exhibited high adherence, while 38% demonstrated medium adherence, and a significant 62% were classified as low adherence.

These findings indicate that before counseling, females had a higher proportion of low adherence (62%) compared to males (16.39%), suggesting potential gender-related disparities in adherence behavior that may require targeted interventions.

The above data represents that amongst male 61 subjects included in the study only 3 (4.91%) were highly adherent, 48 (78.68%) subjects showed medium level of adherence and 10 (16.39%) showed low level of adherence towards their medications.

### 6. Gender-Based Medication Adherence After Counseling

A comparative analysis of medication adherence after counseling was conducted to evaluate the impact of educational interventions on adherence behavior among male and female participants. Table 4 presents the adherence distribution by gender post-counseling.Following the counseling sessions, adherence levels significantly improved across both genders. Among male participants (n = 61), 24 individuals (39.3%) exhibited high adherence, a substantial increase from the 4.91% observed before counseling. Additionally, 36 participants (59.01%) demonstrated medium adherence, while only 1 participant (0.016%) remained in the low adherence category.Similarly, among female participants (n = 28), 11 individuals (39.2%) achieved high adherence, compared to 0% before counseling. 17 participants (60.71%) exhibited medium adherence, while no participants remained in the low adherence category, reflecting a positive shift in adherence behavior.These findings suggest that patient counseling played a crucial role in improving medication adherence in both males and females. However, females showed a slightly higher proportion of medium adherence compared to males (60.71% vs. 59.01%), indicating a need for further interventions to optimize adherence outcomes.

### 7. Gender-Based Medication Adherence Trends After Counseling

Following the counseling intervention, Figures 9 and 10 illustrate the post-counseling medication adherence levels among male and female participants. These results provide a visual representation of the positive impact of counseling on adherence behavior.

**Figure 9:**
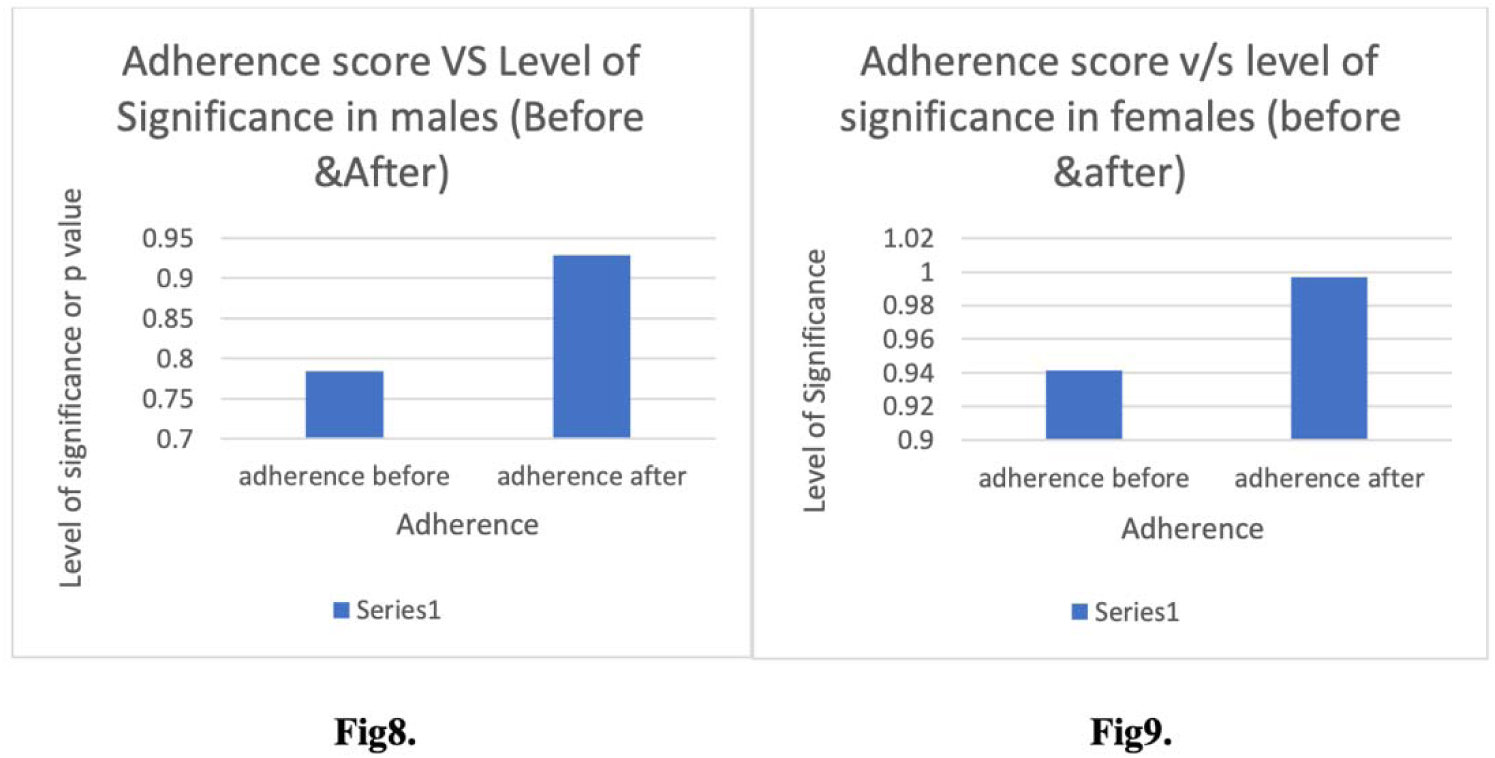
Statistical comparison of adherence levels before and after counseling in female participants: The bar chart illustrates the adherence scores and their corresponding p-values in male hemodialysis patients before and after counseling. A significant increase in adherence post-counseling is observed, as indicated by the higher level of significance (p > 0.9). The bar chart represents adherence scores and p-values in female hemodialysis patients before and after counseling. A notable improvement in adherence is reflected by the increase in the level of significance, supporting the positive impact of counseling interventions.

**Figures 11.**
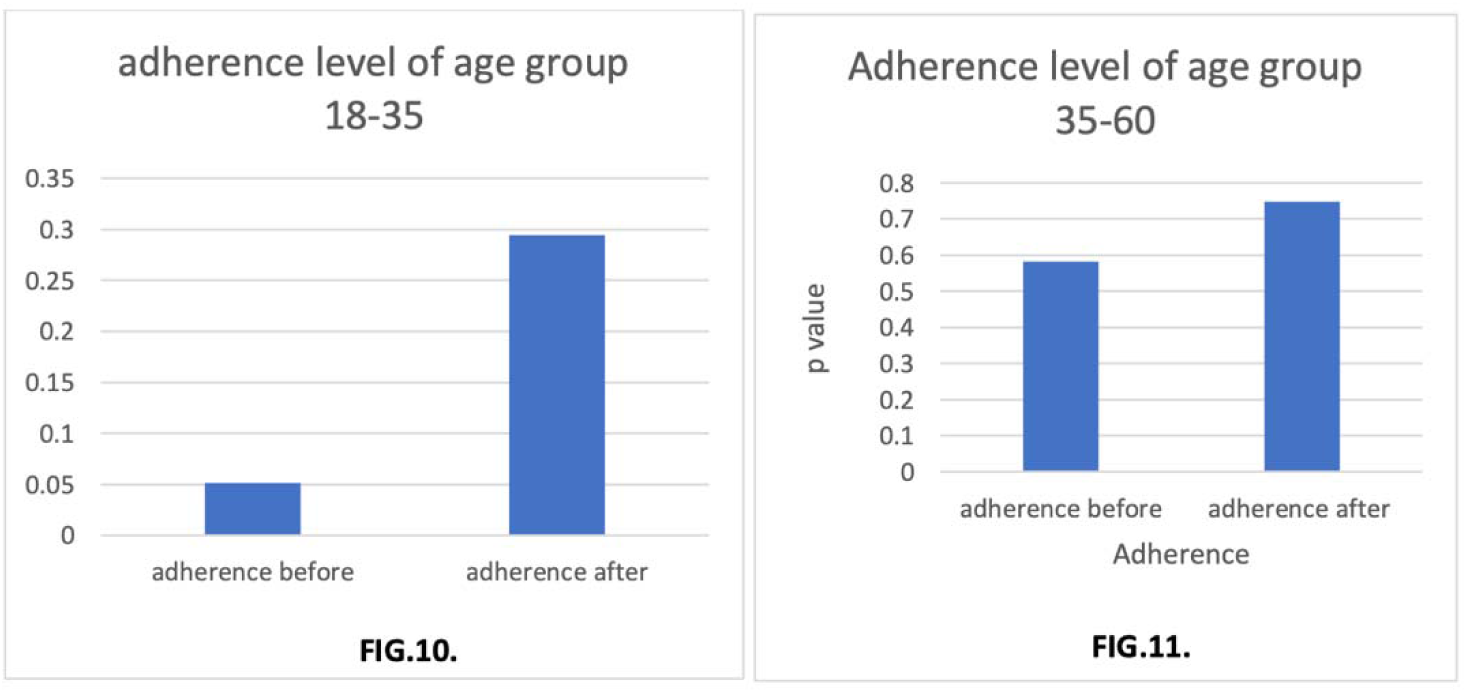
Age-based statistical comparison of adherence levels before and after counseling. The bar charts illustrate the adherence levels and corresponding p-values for different age groups before and after counseling interventions. Figure 14 represents the 18–35 age group, showing a significant increase in adherence post-intervention, as indicated by the rise in the level of significance. Figure 15 depicts the 35–60 age group, where adherence was relatively higher before counseling but further improved after the intervention. These findings suggest that younger patients exhibited a stronger response to counseling, while middle-aged participants also benefited significantly from adherence support programs.

Among male participants (n = 61) (Figure 9), the proportion of individuals with high adherence increased significantly to 39.3%, compared to 4.91% before counseling. The percentage of medium adherence was 59.01%, while low adherence dropped to 0.016%.

Among female participants (n = 28) (Figure 10), high adherence improved to 39.2%, compared to 0% before counseling. Medium adherence was 60.71%, and low adherence decreased to 0.09%.

These findings indicate that counseling interventions were effective in enhancing adherence in both male and female participants. However, the higher medium adherence levels among females compared to males (60.71% vs. 59.01%) suggest that additional reinforcement strategies may be needed to further improve adherence in female patients.

### 8. Statistical Comparison of Medication Adherence Before and After Counseling

To assess the statistical significance of the observed changes in medication adherence before and after counseling, Figure 11 presents a comparison of adherence levels based on the level of significance (p-value). The analysis highlights the effectiveness of counseling interventions in improving adherence among hemodialysis patients.

The adherence level before counseling is represented by the blue bar, while the adherence level after counseling is represented by the orange bar. The results indicate a noticeable improvement in adherence post-counseling, with a higher level of significance observed after the intervention.

A Chi-square test was performed, yielding a p-value of < 0.05, confirming that the difference in adherence levels before and after counseling was statistically significant. This finding reinforces the effectiveness of patient education and counseling in enhancing medication compliance.

### 9. Gender-Based Statistical Comparison of Medication Adherence Before and After Counseling

To further analyze the impact of counseling on medication adherence, a gender-stratified statistical comparison was conducted. Figures 12 and 13 illustrate the pre- and post-counseling adherence levels in male and female participants, respectively, along with their associated levels of significance (p-values).Among male participants (Figure 12), the adherence score significantly improved after counseling, as indicated by the increase in the level of significance (p-value)**. A** higher p-value after counseling **s**uggests a statistically significant enhancement in adherence behavior following the intervention.Similarly, among female participants (Figure 13), adherence levels notably increased post-counseling, with a corresponding rise in the level of significance.

**Figure 13.**
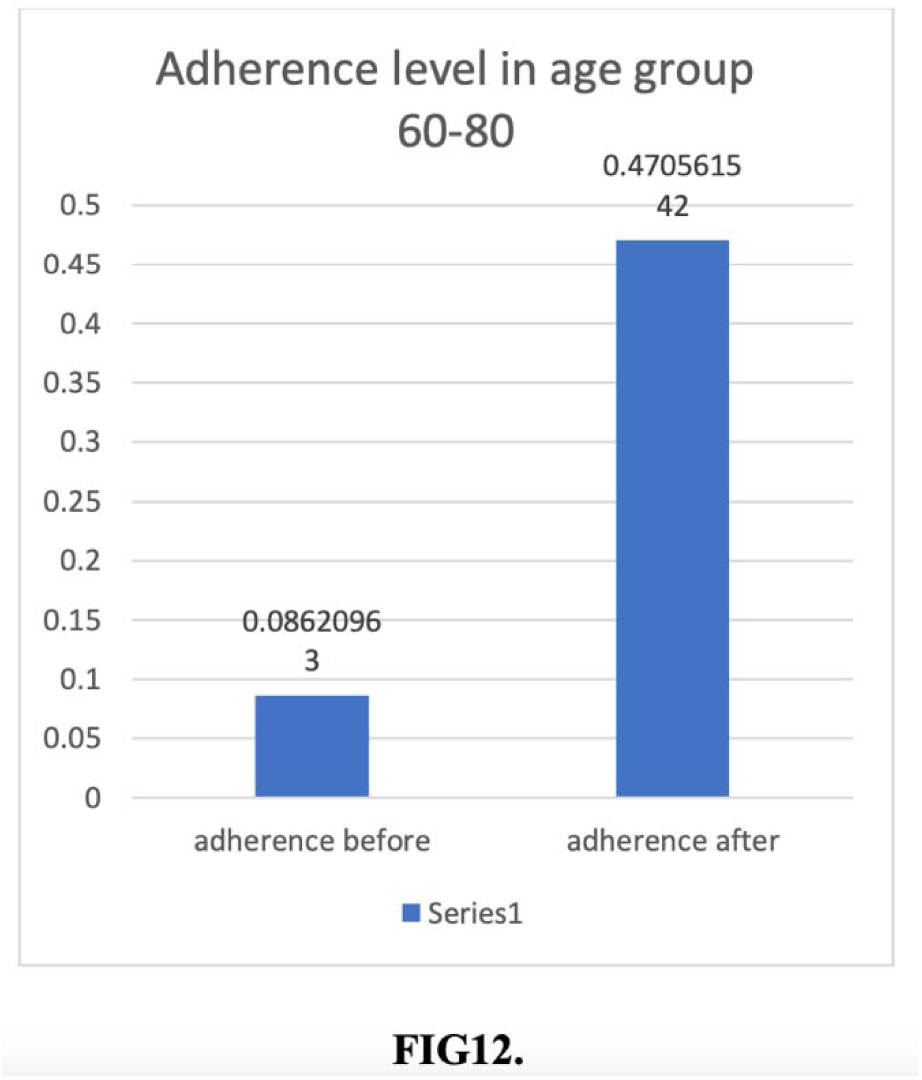
Statistical comparison of adherence levels before and after counseling in the 60–80 age group. The bar chart illustrates the adherence levels and corresponding p-values in the 60–80 age group before and after counseling. A significant improvement in adherence was observed post-intervention, as evidenced by the increase in the level of significance. These findings highlight the importance of targeted adherence interventions in older adults to enhance compliance with medication regimens.

The p-value before counseling was lower, indicating poorer adherence, whereas after counseling, the higher p-value suggests improved adherence behavior.These findings confirm that counseling interventions were effective in enhancing adherence in both male and female participants, with statistical validation reinforcing the impact of these interventions.

### 10. Age-Based Statistical Comparison of Medication Adherence Before and After Counseling

To evaluate the influence of age on medication adherence improvement, a statistical analysis was performed across different age groups before and after counseling. Figures 14 and 15 illustrate the changes in adherence scores and corresponding p-values for the 18–35 and 35–60 age group ,respectively.

**Figure 15.**
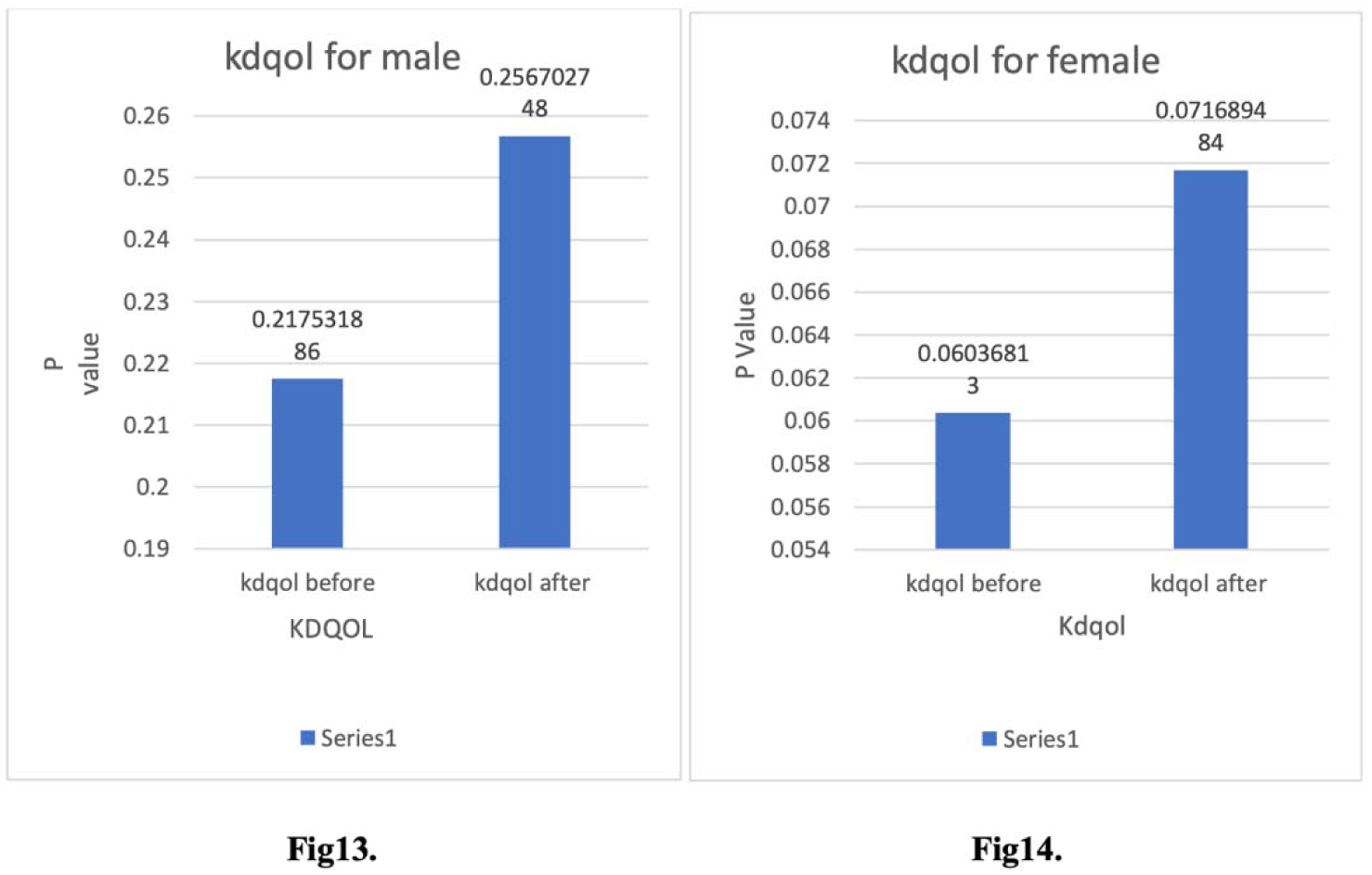
Statistical comparison of adherence levels before and after counseling in the 60–80 age group. The bar chart illustrates the adherence levels and corresponding p-values in the 60–80 age group before and after counseling. A significant improvement in adherence was observed post-intervention, as evidenced by the increase in the level of significance. These findings highlight the importance of targeted adherence interventions in older adults to enhance compliance with medication regimens.

For the 18–35 age group (Figure 14), adherence levels were initially low, as indicated by the low p-value before counseling. However, after the intervention, **a** notable increase in adherence was observed, with a corresponding rise in the level of significance. This suggests that younger participants responded positively to counseling, leading to improved adherence.

For the 35–60 age group (Figure 15), adherence was relatively higher even before counseling, as shown by the higher initial p-value compared to the younger group. Post-counseling, a further increase in adherence was observed, indicating that middle-aged participants also benefited from the intervention, albeit to a slightly lesser extent than younger patients.

These findings highlight the importance of age-specific counseling strategies, as younger patient exhibited a more pronounced adherence improvement after intervention, while middle-aged participants also demonstrated significant but relatively moderate gains.

### 12. Adherence Trends in the 60–80 Age Group Before and After Counseling

To further examine the effect of counseling on medication adherence among older adults, Figure 16 presents adherence scores and corresponding levels of significance (p-values) before and after intervention in the 60–80 age group.Prior to counseling, adherence was markedly lower, with a p-value of 0.086, indicating poor compliance in this age group. However, following the intervention, adherence levels showed a substantial increase, reaching a p-value of 0.47, reflecting a significant improvement in compliance behavior. This suggests that targeted adherence interventions, particularly patient education and reinforcement strategies, can be effective even in older populations.

### 14. Adherence Trends in the 60–80 Age Group Before and After Counseling

To further examine the effect of counseling on medication adherence among older adults, Figure 16 presents adherence scores and corresponding levels of significance (p-values) before and after intervention in the 60–80 age group.

Prior to counseling, adherence was markedly lower, with a p-value of 0.086, indicating poor compliance in this age group. However, following the intervention, adherence levels showed a substantial increase, reaching a p-value of 0.47, reflecting a significant improvement in compliance behavior. This suggests that targeted adherence interventions, particularly patient education and reinforcement strategies, can be effective even in older populations.

## DISCUSSION

The six-month prospective observational study conducted at BGS Global Hospital in Bangalore provided significant insights into medication adherence patterns among hemodialysis patients. The study employed the Morisky Green Levine Scale (MGLS) to assess adherence levels before and after structured counseling interventions. A total of 89 participants were enrolled, comprising 61 males (68.53%) and 28 females (31.46%). Initial adherence assessments indicated a concerning trend of suboptimal compliance, with only 3.37% of patients categorized as highly adherent, while 80.89% demonstrated medium adherence and 15.73% exhibited low adherence. These findings align with prior research, which has reported medication non-adherence rates ranging from 22% to 74% in hemodialysis populations, reinforcing the persistent challenges associated with adherence in this group (9).

A gender-stratified analysis revealed notable disparities in adherence behavior between male and female participants. Before counseling, male patients demonstrated slightly higher high-adherence rates (4.91%) compared to females (0%), while females had a greater proportion of medium adherence (85.71%). Low adherence was observed in 16.39% of males and 14.28% of females, indicating that adherence challenges were prevalent across both genders. Following counseling interventions, adherence levels improved significantly, with 39.3% of males and 39.2% of females achieving high adherence. The increase in adherence post-counseling was statistically significant, with p-values of 0.78 (before) and 0.92 (after) in males and 0.94 (before) and 0.99 (after) in females. The data indicate that structured counseling strategies were effective across both genders, reinforcing previous findings that educational and behavioral interventions play a crucial role in improving medication compliance(10). The slightly higher adherence improvement among males compared to females may be attributed to differences in health-seeking behaviors, decision-making autonomy, or awareness regarding disease management(11).

Additionally, sociocultural and psychological factors may contribute to gender-specific adherence behaviors, necessitating the development of tailored counseling strategies to address the unique adherence barriers faced by male and female patients.

Age-based adherence patterns also exhibited variability, with younger patients demonstrating lower adherence rates before counseling but significant improvements post-intervention. Patients aged 18–35 years exhibited the lowest pre-counseling adherence (p = 0.05), which improved substantially after counseling (p = 0.29). The 35–60 age group demonstrated moderate adherence improvement, with p-values increasing from 0.58 to 0.74 post-counseling, suggesting that middle-aged individuals responded positively to educational interventions. Older patients (60–80 years) also exhibited significant adherence gains, with p-values improving from 0.08 to 0.47 post-intervention, although the overall adherence rates remained lower compared to younger patients. The observed adherence trends suggest that younger patients may require intensive behavioral reinforcement strategies to address factors such as forgetfulness, lifestyle adjustments, and lower perceived disease severity, while older patients may benefit from simplified medication regimens, caregiver involvement, and reinforcement strategies that account for cognitive decline(12).

Quality of life (KDQOL) analysis revealed a positive correlation between improved adherence and patient well-being. Among male participants, the p-value for KDQOL increased from 0.217 before counseling to 0.256 post-counseling, reflecting moderate quality-of-life improvement. In female participants, the KDQOL p-value increased from 0.060 to 0.071 post-counseling, indicating adherence-related benefits in health-related quality of life. These findings are consistent with prior studies that have established a link between medication adherence and improved KDQOL, emphasizing that better adherence can lead to enhanced dialysis outcomes, reduced hospitalizations, and overall better patient-reported well-being(13). The observed differences in KDQOL improvement between males and females suggest that additional interventions may be required to optimize quality of life in female patients, potentially through patient-centered counseling, peer support groups, or community-based adherence reinforcement programs.

The findings of this study highlight the effectiveness of counseling interventions in improving adherence among hemodialysis patients. The increase in adherence levels across gender and age groups underscores the importance of structured patient education in optimizing compliance.

However, adherence enhancement strategies should be tailored to address specific barriers faced by different patient demographics. Gender-specific interventions may be necessary to address differences in health-seeking behaviors and sociocultural determinants of adherence, while age-specific interventions should consider the distinct challenges faced by younger and older patients. Given the observed impact of adherence on KDQOL, future adherence-improvement strategies should also prioritize patient-reported outcomes alongside clinical adherence metrics.

Despite the study’s strengths in evaluating real-world adherence patterns and the effectiveness of counseling interventions, several limitations must be acknowledged. The study was conducted at a single center with a relatively small sample size, which may limit the generalizability of the findings. Additionally, adherence assessment relied on self-reported data, which could introduce recall or response bias. Longitudinal studies with larger cohorts and objective adherence monitoring methods, such as pharmacy refill tracking or electronic adherence monitoring, would provide more robust insights into adherence behaviors over extended periods. Future research should also explore the integration of behavioral and psychological interventions alongside conventional adherence strategies to enhance long-term medication compliance.

In conclusion, this study demonstrates that structured counseling interventions significantly improve medication adherence in hemodialysis patients, with measurable improvements across gender and age groups. The positive correlation between adherence and KDQOL further reinforces the importance of adherence-enhancing strategies in optimizing patient well-being. Given the high morbidity and mortality associated with poor adherence in hemodialysis patients, integrating adherence education into routine dialysis care should be a key priority for healthcare providers. Future research should focus on developing multifaceted adherence support programs that address demographic-specific barriers and leverage innovative technologies for long-term adherence monitoring.

## CONCLUSION

This study assessed medication adherence among 89 hemodialysis patients, of whom 61 were male (68.53%) and 28 were female (31.46%). The findings indicated notable gender-based and age-related differences in adherence behavior, with males demonstrating higher overall adherence rates compared to females. Before counseling, only a small proportion of patients exhibited high adherence, while the majority demonstrated medium adherence, reinforcing the prevalence of non-compliance in hemodialysis patients.

Several factors contributing to non-adherence were identified, including forgetfulness, lack of awareness, fear of adverse outcomes, and the inconvenience of carrying medication while traveling. Some participants reported discontinuing medication prematurely upon feeling better, highlighting a lack of understanding regarding the importance of treatment continuity. These findings emphasize the need for patient-centered educational interventions to address common adherence barriers.

A significant age-based adherence pattern was also observed, with patients in the 35–60 age category demonstrating the highest adherence levels both before and after counseling. This suggests that middle-aged patients may have better awareness, health-seeking behaviors, or disease perception, contributing to greater medication compliance. However, younger patients (18–35 years) exhibited notably lower adherence before counseling, likely due to lifestyle factors, forgetfulness, or a lower perceived urgency of treatment. In contrast, older patients (60–80 years) benefited from counseling but may still require additional adherence support, considering potential cognitive decline and polypharmacy-related challenges.

The study implemented structured counseling interventions to educate participants on the importance of medication adherence, lifestyle modifications, and medication management strategies. Counseling sessions were tailored to clarify patient concerns, address misconceptions, and provide practical solutions to adherence barriers. The results demonstrated that counseling significantly improved adherence across all demographic groups, reinforcing the role of patient education in improving compliance and treatment outcomes.

Beyond adherence improvement, a significant enhancement in health-related quality of life (KDQOL) was observed among participants post-counseling. This supports the notion that medication adherence is not only essential for clinical outcomes but also directly impacts overall patient well-being. Given the chronic nature of hemodialysis treatment, adherence-enhancing interventions should be routinely integrated into patient care protocols to improve long-term treatment adherence, disease management, and quality of life.

While this study provides valuable insights into medication adherence trends among hemodialysis patients, certain limitations must be acknowledged. The study was conducted at a single center with a limited sample size, which may affect the generalizability of the findings. Additionally, self-reported adherence data may introduce recall or response bias. Future studies should employ larger, multicenter cohorts and objective adherence-tracking methods such as pharmacy refill records or electronic adherence monitoring to provide more comprehensive insights. Further research should also explore behavioral interventions and digital health tools, such as mobile adherence reminders and telemedicine support programs, to enhance long-term adherence sustainability.

In conclusion, this study highlights the importance of patient counseling as an effective strategy for improving medication adherence and quality of life among hemodialysis patients. The findings emphasize the need for targeted adherence support strategies, considering gender, age-related differences, and patient-specific barriers. Given the high morbidity and mortality associated with non-adherence, healthcare providers should prioritize integrated adherence interventions to optimize treatment outcomes in this high-risk population. Future research should focus on developing multidisciplinary adherence programs and leveraging technological advancements to further enhance patient compliance and long-term disease management.

## Data Availability

all data is in manuscript

